# “Breaking the silence”: Sexual victimisation in an old age psychiatry patient population in Flanders

**DOI:** 10.1101/2021.02.03.21251063

**Authors:** Anne Nobels, Ines Keygnaert, Egon Robert, Christophe Vandeviver, An Haekens, Lieve Lemey, Marieke Strobbe, Nele Van Den Noortgate, Gilbert M.D. Lemmens

**Affiliations:** International Centre for Reproductive Health, Department of Public Health and Primary Care, Ghent University, Ghent, Belgium; Department of Criminology, Criminal Law and Social Law, Ghent University, Ghent, Belgium; Research Foundation—Flanders (FWO), Brussels, Belgium; Psychiatric Hospital Alexianen Zorggroep Tienen, Tienen, Belgium; Department of Psychiatry and Psychosomatic Medicine, AZ Sint-Jan Bruges-Ostend AV, Bruges, Belgium; Psychiatric Hospital Karus, Melle, Belgium; Department of Geriatric Medicine, Ghent University Hospital, Ghent, Belgium; Department of Psychiatry, Ghent University Hospital, Ghent, Belgium; Department of Head and Skin, Psychiatry and Medical Psychology, Ghent University, Ghent, Belgium

**Keywords:** Sexual abuse, sexual assault, elder abuse and neglect, geriatric psychiatry

## Abstract

**Background:** Sexual violence (SV) is linked to mental health problems in adulthood and old age. However, the extent of sexual victimisation in old age psychiatry patients is unknown. Due to insufficient communication skills in both patients and healthcare workers, assessing SV in old age psychiatry patients is challenging.

**Methods:** Between July 2019 and March 2020, 100 patients at three old age psychiatry wards across Flanders participated in a face-to-face structured interview receiving inpatient treatment. The participation rate was 58%. We applied the WHO definition of SV, encompassing sexual harassment, sexual abuse with physical contact without penetration, and (attempted) rape.

**Outcomes:** In 57% of patients (65% F, 42% M) SV occurred during their lifetime and 7% (6% F, 9% M) experienced SV in the past 12-months. Half of the victims disclosed their SV experience for the first time during the interview. Only two victims had disclosed SV to a mental health care professional before.

**Interpretation:** Sexual victimisation appears to be common in old age psychiatry patients, yet it remains largely undetected. Although victims did reveal SV during a face-to-face interview to a trained interviewer, they do not seem to spontaneously disclose their experiences to mental health care professionals. In order to provide tailored care for older SV victims, professionals urgently need capacity building through training, screening tools and care procedures.

## 1. Introduction

Increasingly, sexual violence (SV)^1^ is considered a public health problem with an important impact on the victim’s mental health.^2^ Research consistently links sexual victimisation to a wide range of mental health conditions in adulthood.^3^ Moreover, people with severe mental illness (SMI) are at substantially increased risk of experiencing SV during their lifetime compared to those who do not suffer from SMI.^4^ Research shows that SV victims experience many barriers for disclosure and as a result do not receive the appropriate (mental health) care.^5^ Therefore, the World Psychiatric Association (WPA) released a “Position Statement on Intimate Partner Violence and SV against Women” in which they petition for routine inquiry on SV to be included in all psychiatric assessments of female patients.^6^

In older adults, SV seems to rarely occur. A recent meta-analysis showed that 0.9% of community-dwelling older adults were sexually victimised in the past 12-months.^7^ A Belgian study found that 6% of older adults have been exposed to SV during their life.^8^ However, due to methodological shortcomings, these numbers are likely to be underestimated.^9^ In old age psychiatry patients the extent of sexual victimisation is still unknown. Since several studies link exposure to adverse life events in childhood, including SV, to internalizing disorders in old age^10,11^, we can only assume that a high proportion of old age psychiatry patients were exposed to SV during their lifetime.

Furthermore, assessing sexual victimisation in old age psychiatry patients may be challenging. Older adults grew up in a time when talking about sexuality and SV was considered taboo. Due to a lack of comprehensive sexual education (CSE) and different legal definitions or ideas on sexual consent, old age psychiatry patients may have different perceptions of SV compared to younger patients.^12^ Moreover, older adults are considered asexual by society.^13^ When internalizing this stereotypical societal image of ‘the asexual older adult’, old age psychiatry patients may not identify themselves as possible SV victims.^14^ Consequently, they were not inclined to disclose sexual victimisation nor to seek medical or psychological help ^14^. In addition, health care workers tend not to address sexual health (SH) and SV proactively with older patients, as they feel it is not a legitimate topic to discuss with this age group and are afraid of offending their patients.^15,16^ Moreover, they seem to lack the appropriate communication skills to adequately deal with SV in later life.^17^

Since 30% of the European population and 25% of the Belgian population is estimated to be 65 years or older by 2060^18^, an increased need for mental health care services for older adults is to be expected. Due to the major impact of SV on mental health, an assessment of sexual victimisation in older psychiatric patients seems necessary in order to provide tailored mental health care in this patient group. To our knowledge, this paper is the first study to assess lifetime and sexual victimisation in the past 12-months, correlates and SV disclosure in an old age psychiatry population. Based on the results, we formulate recommendations for clinical practice and future research.

## 2. Methods

### 2.1 Sample selection

Between the 9^th^ of July 2019 and the 12^th^ of March 2020, all consecutive patients receiving inpatient treatment at three old age psychiatry wards in three different cities (Bruges, Ghent and Tienen) across Flanders were invited to participate in the study. Based on the 10% SV prevalence rates found in previous studies done in the Belgian population^19^, we calculated a target sample of 138 participants (see Appendix 1). Unfortunately, the COVID-19 pandemic and associated lockdown measures forced us to stop data collection before the target sample size could be reached.

In order to assure comparability with a SV prevalence study in Belgian older adults conducted at the same time, only patients of 70 years and older could participate. Only patients receiving inpatient treatment were included. Patients needed to speak Dutch, French or English and have sufficient cognitive ability to complete the interview. Cognitive status was not formally assessed, but was evaluated based on the ability to maintain attention during the interview and the consistency of the participant’s answer, by means of a control question comparing the participant’s birth year and age. Patients suffering from an acute psychotic episode were excluded from participating, but could participate after remission of psychotic symptoms.

During the data collection period, 210 individual patients of 70 years and older were admitted at the three old age psychiatry wards. Of those patients 14 left the hospital before they could be contacted by our interviewers, 25 suffered from psychotic symptoms during the entire length of their hospitalization and 12 patients were not able to participate because of severe cognitive impairment. Of the 173 eligible patients, 101 agreed to participate. One interview was stopped prematurely due to the participant displaying cognitive problems and was not included in the analysis. Four interviews were stopped prematurely at the request of the participants, but were included in parts of the analysis. Ultimately 100 interviews were included in the analysis, which provides us with a response rate of 58%.

### 2.2 Definitions and measures

SV was defined according to the WHO definition as “Every sexual act directed against a person’s will, by any person regardless of their relationship to the victim, in any setting”.^1^ It includes different forms of sexual harassment without physical contact, sexual abuse with physical contact but without penetration and (attempted) rape.^1^ This definition was expanded to include sexual neglect, as a result of recent insights in the field of SV in older adults.^9^ The questionnaire consisted of 12 modules and its design aimed to maximize SV disclosure. To this extent, the questionnaire started with less sensitive topics and built up towards the SV modules. Lifetime and past 12-months SV experiences were assessed using behaviourally specific questions (BSQ) based on the Sexual Experience Survey (SES)^20^, the Sexual Aggression and Victimization Scale (SAV-S)^19^, and the Senperforto questionnaire^21^ which were adapted to the Belgian social and legal context.^22^ The use of BSQ is recommended in order to provide reliable estimates of both female and male sexual victimisation.^23^ Mental health was measured using validated scales: the Patient Health Questionnaire (PHQ)-9 for depression^24^ (Cronbach alfa (α) =0·765), the General Anxiety Disorder (GAD)-7 for anxiety^25^ (α=0·850), the Primary Care PTSD Screen for DSM-5 (PC-PTSD-5) for PTSD^26^ (α=0·591), the Alcohol Use Disorder Identification Test-Short version (AUDIT-C) for hazardous alcohol use^27^ (α=0·784), and the Brief Resilience Scale (BRS)^28^ to assess resilience (α=0·773). In addition, participants were asked about sedative use, drug use, self-harm and suicide attempts, both during their lifetime and in the past 12-months.

### 2.3 Ethical considerations

The study was conducted according to the WHO ethical and safety recommendations for SV research^29^ and received ethical approval from the ethical committee of Ghent University/University Hospital and the ethical committees of the three participating hospitals (Belgian registration number: B67021939483). As recommended in SV research^29^, the study was presented as “Survey on Health, Sexuality and Well-being”. All patients gave their informed consent before participating. Interviews were conducted in private in a separate room in the hospital by a psychiatry resident and three master’s students of medicine. Interviewers received a three day training on SV, interview techniques and handling confidential information prior to the fieldwork. During the fieldwork, interviewers received close guidance from the research team. In line with current EU privacy legislation, all data were pseudonymised. Documents containing personal data were kept separately from the answers to the questionnaire. Only the researchers working on the project had access to the personal data. In case of an acutely dangerous situation (e.g. ongoing (sexual) violence), a multi-step safety procedure was in place in accordance with article 458 of Belgian criminal law. At the end of each interview, participants received a brochure containing the contact details of several helplines.

### 2.4 Statistical analysis

Statistical analysis was performed using SPSS Statistics 25. Descriptive statistics were used to illustrate the sociodemographic characteristics, mental health status and sexual victimisation of the study sample. SV variables were grouped into hands-off (eight items) and hands-on SV (nine items), the latter being further grouped into sexual abuse (four items) and attempted or completed rape (five items). For the purpose of the analysis the item measuring sexual neglect (touching in care) was grouped under sexual abuse. We created dichotomous variables out of all items in order to assess lifetime and past 12-months sexual victimisation. A detailed overview of the SV outcome measures can be found in Appendix 2.

A number of demographical, socioeconomical and variables related to the patients’ sexuality and mental health status were included in the correlation analysis. Because of the overlap between hands-off and hands-on victimisation in our sample, no correlation analysis was performed for hands-off and hands-on SV separately. In the bivariate analysis, categorical variables were analysed dichotomously using the Pearson’s chi-square test. The Student’s T-test was used for continuous variables, after checking for normality. Multivariate binary logistic regression was performed to identify predictors of lifetime SV. All variables used in the bivariate testing were entered into the model using backward binary regression analysis. P-values, odds ratios (OR) and 95% confidence intervals (CI) are presented.

## 3. Results

### 3.1 Study population characteristics

Table 1 describes the sociodemographic characteristics of the study population. The mean age was 78 years (SD: 6.16yrs, range: 70-94yrs), 66% was female, 97% was born in Belgium and 25% completed higher education. Over 50% had a partner, 43% did not have a partner and 40% lived alone. Considering sexual orientation, 18% labelled themselves as non-heterosexual.

**Table 1.** Sociodemographic characteristics of the study population (n = 100)

| Variable |  | n & % |
| --- | --- | --- |
| Sex | Female | 66 |
|  | Male | 34 |
| Age<br>(mean 78y) | 70-79y | 62 |
|  | 80-89y | 33 |
|  | 90- 99y | 5 |
| Country of origin | Belgium | 97 |
|  | Other | 3 |
| Education level | Primary education | 33 |
|  | Secondary education | 42 |
|  | Higher education | 25 |
| Sexual orientation<sup>a</sup> | Heterosexual | 82 |
|  | Non-heterosexual | 18 |
| Relationship status | Living together with partner | 52 |
|  | Relationship, but living apart | 5 |
|  | No relationship/ partner | 43 |
| Living situation | Alone | 40 |
|  | Not alone | 60 |

<sup>a</sup>This group contains participants who labelled themselves as: homosexual, bisexual, pansexual, asexual or other. In this last group, several participants labelled themselves as “normal”. Since it was not clear whether they had difficulties understanding the different terms defining sexual orientation or whether they indeed labelled their sexual orientation as “other”, we however decided to classify these participants as non-heterosexual based on their answer.

Concerning mental health status (see Table 2), 42·8% reported moderate to severe depressive symptoms, 32·7% moderate to severe anxiety symptoms, 25·0% showed symptoms of PTSD and 23·0% reported hazardous alcohol use. In the past 12-months, 78·0% of the study population used sedatives, 10·3% attempted suicide and 1·0% reported self-harm.

**Table 2.** Mental health status of the study population, presented by sex.

| Item | Scale | $\alpha$ | Outcome | Men<br>n (%) | Women<br>n (%) | Total<br>n (%) |
| --- | --- | --- | --- | --- | --- | --- |
| Depression (n = 98) | PHQ-9 | 0.765 | Mild | 10 (29.4) | 21 (32.8) | 31 (31.6) |
|  |  |  | Moderate | 9 (26.5) | 9 (14.1) | 18 (18.4) |
|  |  |  | Moderately severe | 4 (11.8) | 14 (21.9) | 18 (18.4) |
|  |  |  | Severe | 1 (2.9) | 5 (7.8) | 6 (6.1) |
| Anxiety (n = 98) | GAD-7 | 0.850 | Mild | 5 (14.7) | 18 (28.1) | 23 (23.5) |
|  |  |  | Moderate | 8 (23.5) | 11 (17.2) | 19 (19.4) |
|  |  |  | Severe | 5 (14.7) | 18 (28.1) | 23 (23.5) |
| PTSD (n = 100) | PC-PTSD-5 | 0.591 | Yes | 10 (29.4) | 15 (22.7) | 25 (25.0) |
| Resilience (n = 99) | BRS | 0.773 | Low | 18 (54.5) | 45 (68.2) | 63 (36.6) |
|  |  |  | Normal | 15 (45.5) | 19 (28.8) | 34 (34.3) |
|  |  |  | High | 0 (0.0) | 2 (3.0) | 2 (2.0) |
| Hazardous alcohol use (n = 100) | AUDIT-C | 0.784 | Yes | 9 (26.4) | 14 (21.2) | 23 (23.0) |
| Sedative use (n = 100) | NA | NA | Lifetime | 28 (82.4) | 61 (92.4) | 89 (89.0) |
|  |  |  | Past 12-months | 25 (73.5) | 53 (80.3) | 78 (78.0) |
| Suicide attempt (n = 97) | NA | NA | Lifetime | 4 (12.1) | 13 (20.3) | 17 (17.5) |
|  |  |  | Past 12-months | 3 (9.1) | 7 (10.9) | 10 (10.3) |
| Self-harm (n = 97) | NA | NA | Lifetime | 0 (0.0) | 2 (3.1) | 2 (2.1) |
|  |  |  | Past 12-months | 0 (0.0) | 1 (1.6) | 1 (1.0) |

$\alpha$ = Cronbach's Alfa, PTSD= Posttraumatic Stress Disorder
Patient Health Questionnaire-9 (PHQ-9): Mild (5-9), Moderate (10-14), Moderately severe (15-19), Severe ( $\geq 20$ )
General Anxiety Disorder-7 (GAD-7): Mild (5-9), Moderate (10-14), Severe ( $\geq 15$ )
Primary Care PTSD Screen for DSM-5 (PC-PTSD-5): Yes ( $\geq 3$ )
Brief Resilience Scale (BRS): Low (1.00-2.99), Normal (3.00-4.30), High (4.31-5.00)
Alcohol Use Disorder Identification Test-Short version (AUDIT-C): Yes ( $\geq 4$ females, $\geq 5$ males)

### 3.2. Sexual victimisation

Lifetime sexual victimisation occurred in 57·3% of our study population, 65·1% of female patients and 42·4% of male patients were sexually victimised during their life. Over half of women and one in three men experienced hands-off SV. Similar numbers were found concerning hands-on SV. One in three participants experienced hands-on SV as a minor (<18 years). One in five female respondents disclosed that they had experienced rape or attempted rape. In the past 12-months, 7·3% experienced at least one form of SV, 3·1% reported hands-off and 4·2% hands-on SV. A more detailed description of the different forms of SV can be found in Table 3.

**Table 3.**
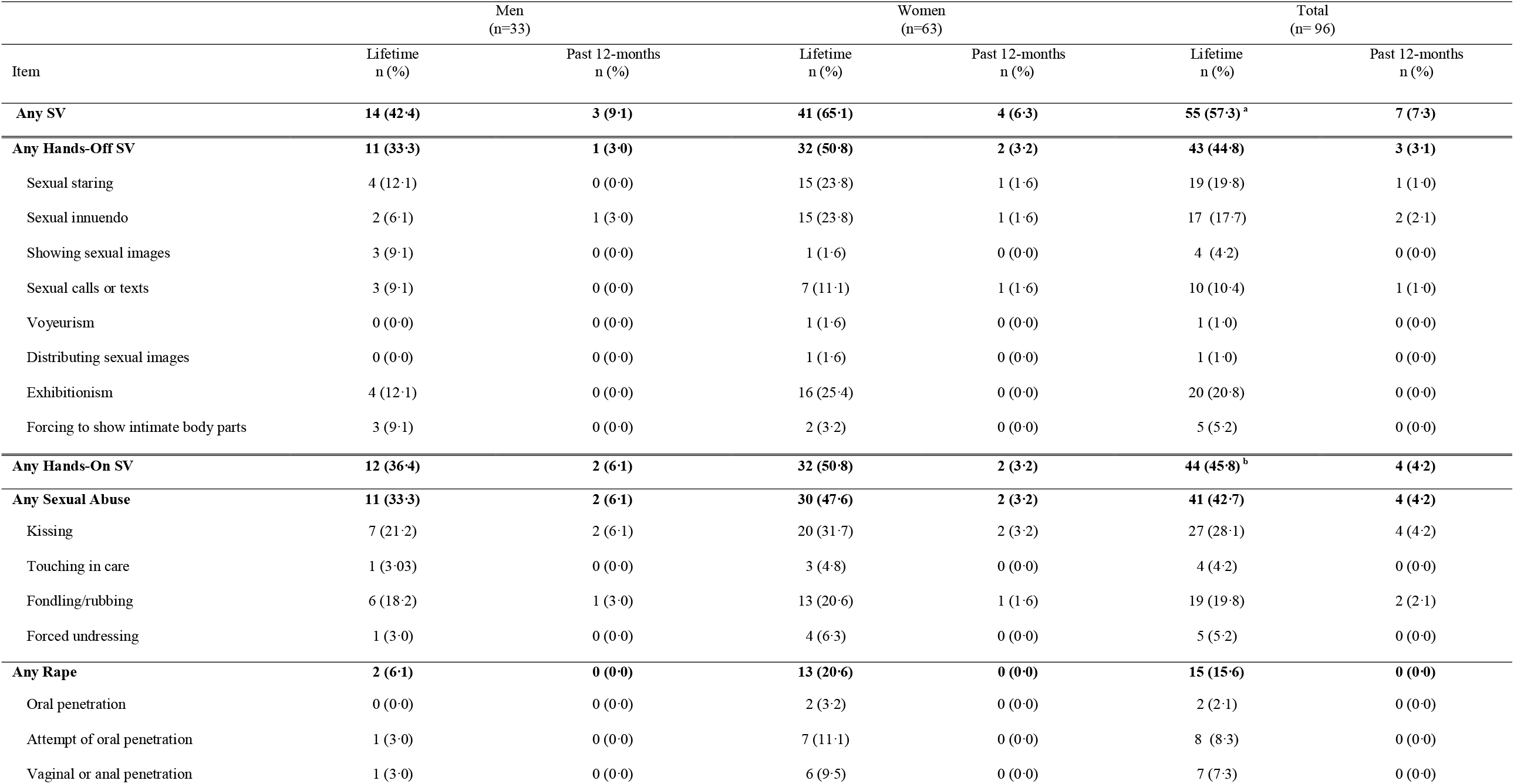
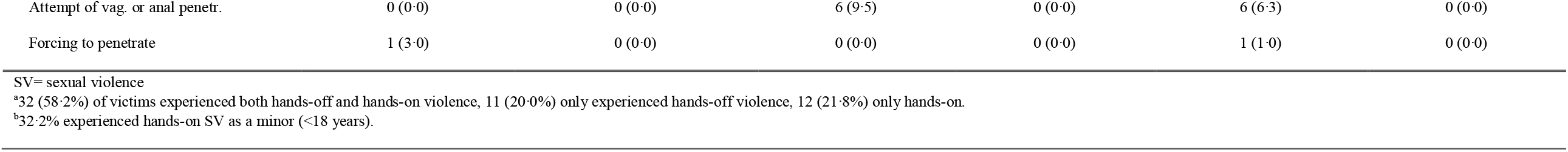
Lifetime and past 12-months sexual victimisation, presented by sex.

### 3.3 Characteristics of victims of sexual violence

Women experienced significantly more SV compared to men (p < 0·05) (see Table 4), except in the past 12-months. Patients without a partner or living separately from their partner and patients with lower levels of education experienced significantly more SV during their lifetime (p < 0·05). We found no difference between heterosexual and non-heterosexual patients regarding sexual victimisation during their lifetime and the past-12 months.

**Table 4.**
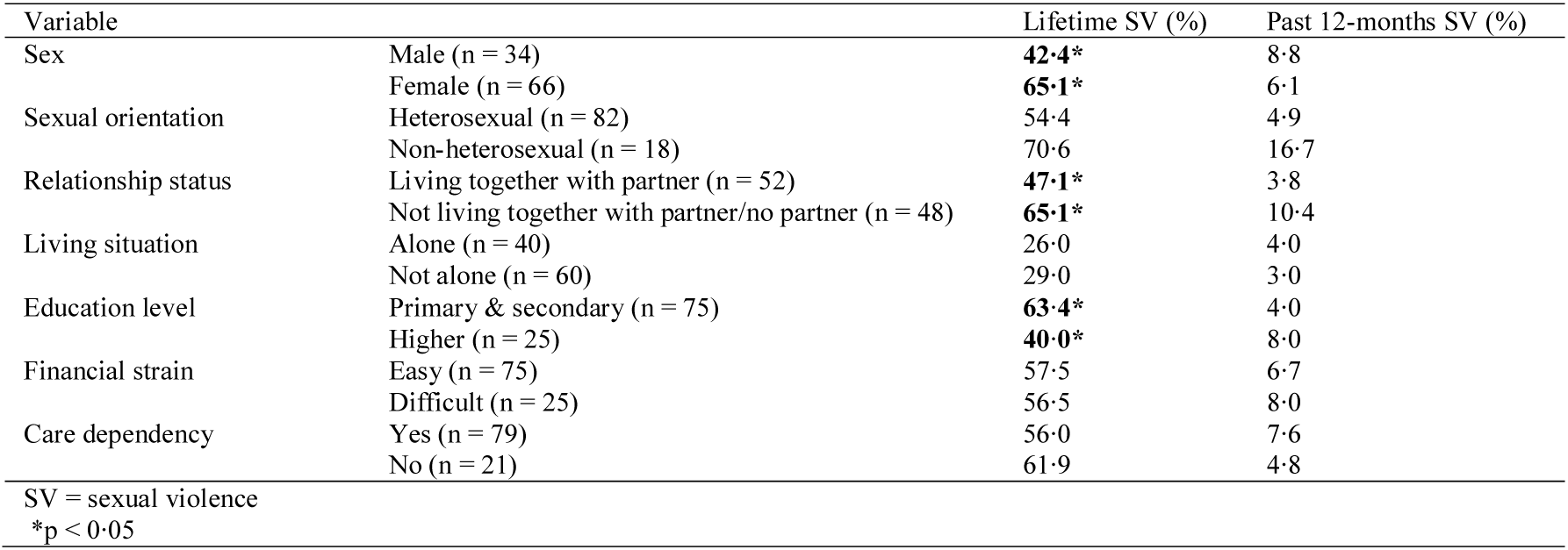
Sociodemographic characteristics and sexual victimisation.

| Variable |  | Lifetime SV (%) | Past 12-months SV (%) |
| --- | --- | --- | --- |
| Sex | Male (n = 34) | **42.4\*** | 8.8 |
|  | Female (n = 66) | **65.1\*** | 6.1 |
| Sexual orientation | Heterosexual (n = 82) | 54.4 | 4.9 |
|  | Non-heterosexual (n = 18) | 70.6 | 16.7 |
| Relationship status | Living together with partner (n = 52) | **47.1\*** | 3.8 |
|  | Not living together with partner/no partner (n = 48) | **65.1\*** | 10.4 |
| Living situation | Alone (n = 40) | 26.0 | 4.0 |
|  | Not alone (n = 60) | 29.0 | 3.0 |
| Education level | Primary & secondary (n = 75) | **63.4\*** | 4.0 |
|  | Higher (n = 25) | **40.0\*** | 8.0 |
| Financial strain | Easy (n = 75) | 57.5 | 6.7 |
|  | Difficult (n = 25) | 56.5 | 8.0 |
| Care dependency | Yes (n = 79) | 56.0 | 7.6 |
|  | No (n = 21) | 61.9 | 4.8 |

SV = sexual violence
\* $p < 0.05$

Although victims of lifetime SV showed a tendency towards more symptoms of depression, anxiety, PTSD, and suicide attempts (see Table 5), these results were not significant. Also, for hazardous alcohol use, sedative use and resilience, no significant differences between victims and non-victims were found. Concerning SV during the past 12-months, victims reported significantly more anxiety symptoms compared to non-victims (p < 0·01). All other mental health outcomes showed no significant results.

**Table 5.** Mental health and sexual victimisation.

| Variable | Lifetime SV | Mean score | Past year SV | Mean score |
| --- | --- | --- | --- | --- |
| Depression (PHQ-9) | Yes | 10.36 | Yes | 12.57 |
|  | No | 7.58 | No | 9.16 |
| Anxiety (GAD-7) | Yes | 9.18 | Yes | **14.29\*** |
|  | No | 7.15 | No | **7.97\*** |
| PTSD (PC-PTSD-5) | Yes | 1.64 | Yes | 2.57 |
|  | No | 1.49 | No | 1.51 |
| Resilience (BRS) | Yes | 2.56 | Yes | 2.19 |
|  | No | 2.79 | No | 2.56 |
| Alcohol (AUDIT C) | Yes | 1.93 | Yes | 1.43 |
|  | No | 2.39 | No | 2.17 |
| Variable | Lifetime SV | Percentage | Past year SV | Percentage |
| Suicide attempt (lifetime) | Yes | 20.0 | Yes | 42.9 |
|  | No | 14.6 | No | 15.6 |
| Sedative use (past 12-months) | Yes | 90.9 | Yes | 85.7 |
|  | No | 85.4 | No | 89.2 |

SV = sexual violence, Patient Health Questionnaire-9 (PHQ-9), General Anxiety Disorder -7 (GAD-7), Primary Care PTSD Screen for DSM-5 (PC-PTSD-5), Brief Resilience Scale (BRS), Alcohol Use Disorder Identification Test-Short version (AUDIT-C)
\*p < 0.05

In the multivariate analysis on lifetime sexual victimisation, only living together with a partner showed a significant association (see Table 6). The PHQ-9 score for depression, care dependency and living together with a partner were identified as the best combined predictors of SV. Because our sample contained fewer than ten victims of SV during the past 12-months, there was no multivariate analysis performed on this group.

**Table 6.**
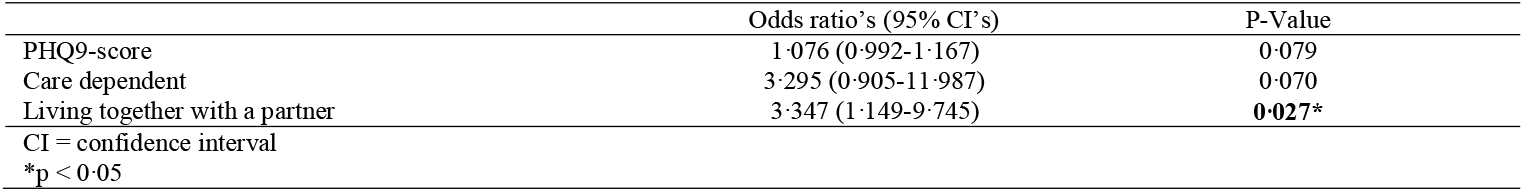
Multivariate analysis for lifetime sexual victimisation.

### 3.4 Assailant characteristics

Over 80% of assailants of lifetime SV were male and of the opposite sex to the victim (see Table 7). Friends (26·4%) were most often identified as assailants, followed by unknown assailants (20·8%), family members (13·3%), an acquaintance (15·1%), authority figures and colleagues/classmates (both 9·4%) and (ex) partners (5·7%).

**Table 7.**
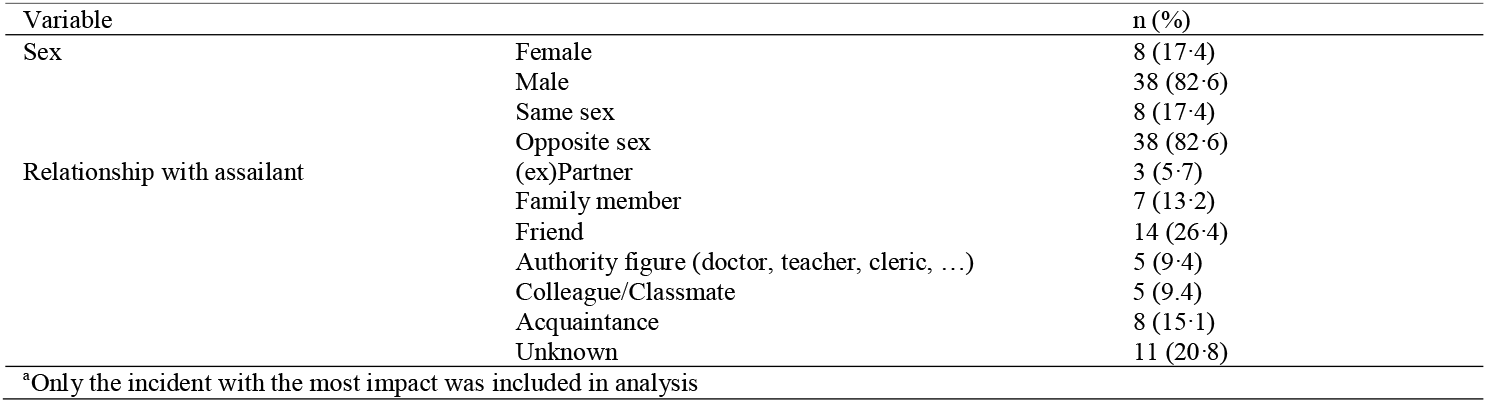
Characteristics of assailants of lifetime sexual violence^a^.

### 3.5 Sexual violence disclosure and interviewers’ experiences

Half of SV victims disclosed their experiences for the first time during the interview (see Table 8). Victims who did disclose their experience, did this most often to their partner (42·3%), followed by parents, family members, acquaintances (all 23·1%) and friends (19·2%). Only 11·5% of victims sought professional help and only two victims had disclosed their experiences to a mental health care professional.

**Table 8.**
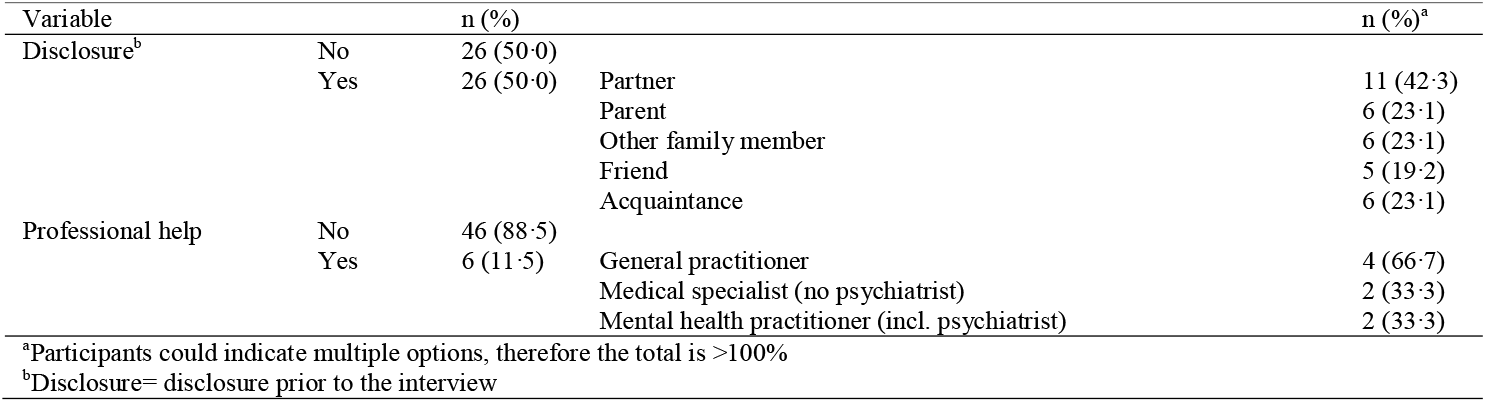
Disclosure & help seeking after sexual victimisation.

During the fieldwork the safety of the respondents was never endangered. One respondent expressed concrete suicidal plans to our interviewer. According to the study’s safety measures this was reported to the patient’s psychiatrist and appropriate care was administered. The interviewers rated the overall experience as rewarding. They received positive feedback from respondents. Participants expressed their appreciation of the study, reporting that they felt heard and treated with respect. They openly discussed sexual health and SV with both same and opposite sex interviewers.

## 4. Discussion

In this paper we present the results of a study on lifetime and past 12-months sexual victimisation in an old age psychiatry population in Flanders. Between the 8^th^ of July 2019 and the 12^th^ of March 2020, we conducted 100 interviews with patients of 70 years and older receiving inpatient treatment at three old age psychiatry wards in three different cities in Flanders.

Our results show that sexual victimisation is highly prevalent within this old age psychiatry population. Over 57% of patients reported being sexually victimised in their lifetime. Over half of women and one in three men experienced hands-off SV. Similar numbers were found concerning hands-on SV. One in five women experienced rape or attempted rape. Exposure to SV in the past 12-months was 7·3%. These numbers are much higher compared to lifetime and past 12-months SV prevalence rates in community-dwelling older adults.^7,8^ Since severe mental illness is associated with an increased risk of SV during lifetime^4^, higher SV rates compared to the general older population could be expected. Nevertheless, compared to previous research in SMI patients, our study shows similar numbers in women, but a higher SV rate in men. A UK study on 303 patients with severe mental illness with a mean age of 40 years, showed that lifetime and past 12-months sexual victimisation in women was respectively 61% and 10%.^4^ In men, lifetime and past 12-months sexual victimisation was 23% and 3% compared to 42% and 9% in our study. A possible explanation could be that male patients are more prone to sexual victimisation after the age of 40 years compared to female patients, leading to an increased rate of lifetime SV in older male psychiatric patients.

However, a recent SV prevalence study in the Belgium population aged 16-69 years contradicts this hypothesis.^30^ Another reason could be the use of BSQ to assess SV in our study. Research has shown that the use of BSQ leads to increased detection of SV in both men and women, but the impact may be even greater in men.^23^ Because of prevailing gender norms and rape myths, male victims often don’t consider themselves as victims of SV. By using BSQ those victims can also indicate their SV experiences, leading to more reliable measures.^23^

The vast majority of assailants (80%) were known to the victim and almost one in five incidents of SV happened within the family of the victim. However, in only 5·7% of the cases the (ex)partner was identified as the assailant. This is much less compared to European studies in younger populations in which over 25% of women report being sexually victimised by their (ex)partner.^2^ It is possible older adults may have a different perception of SV than younger generations, as spousal rape was only criminalised at the end of the 20th century in most European countries.^12^ Although the use of BSQ is supposed to measure sexual victimisation independently of societal norms, it is possible that the difference in perceptions around SV within an intimate partner relationship could have influenced disclosure of SV in this study population and consequently, lead to an underestimation of sexual victimisation rates.

Given the high numbers for sexual victimisation, our study confirms the association between sexual victimisation and mental health problems later in life.^3^ While current international guidelines on detection and care of SV by mental health care professionals focus only on female patients, our study shows that male patients should not be ignored when it comes to SV.^6^ In our sample, female patients seemed to have experienced significantly more SV during their lifetime compared to male patients, but in the multivariate analysis, female sex did not seem to be a predictor of lifetime sexual victimisation.

The majority of patients had never sought medical or psychological help regarding their SV experiences and only two patients had disclosed their experiences to a mental health care professional. This confirms the findings of previous research indicating that victims experience many barriers regarding SV disclosure.^5^ Both victims and health care workers seem to apply the “speech is silver, but silence is golden” principle when it comes to sexual victimisation in later life. Health care professionals are particularly worried that opening Pandora’s box of sexual victimisation will break down the victims’ defence mechanisms and that they will feel shocked and helpless when confronted with SV disclosure by older victims.^17^ In our study we took several measures to ensure patients felt comfortable revealing SV during a face-to-face interview. To maximize disclosure the questionnaire built up towards the SV modules. Furthermore, we carefully selected our interviewers and provided them with specialized training and close support during the data collection period. Interviewers were trained to create a bond of trust with the participants by allowing a certain deviation of topic in order to show interest in the participants’ life. As a result, 50% of victims disclosed their SV experiences for the first time during the interview. Many participants reported that they felt treated respectfully and were grateful to be able to share their story. This indicates that, in spite of the taboo on SV^12^ and the image of the ‘asexual older adult’^13^, older psychiatric patients are willing to “break the silence” and share their experiences of sexual victimisation if they are asked about it. Moreover, the fear of health care workers of offending their patients when talking about SH and SV proves to be untrue.^15,16^ Based on our experiences, we believe that incorporating a routine inquiry about sexual victimisation, as proposed by the WPA^6^, should become the gold standard of care for both female and male old age psychiatry patients. Future research should focus on the necessary conditions and skills to make this possible. In addition, to better detect signs, prevent, mitigate and respond to SV in older patients, professionals urgently need capacity building through training, detection tools and care procedures.

The main limitations of our study are the non-probabilistic sampling method and the specific geographical region of Flanders, which limit the generalisability of our findings. Also, the relatively small sample size reduces the power of our analysis. Nevertheless, this study can be regarded as a first step towards a better understanding of the magnitude, nature and impact of SV within the old age psychiatry population. Based on our results we believe future research could focus on the impact of SV disclosure in later life and psychotherapeutic and psychopharmacological interventions for trauma related disorders as a result of SV in old age.

## 5. Conclusion

Sexual victimisation appears to be common in both female and male old age psychiatry patients, yet it remains largely undetected. Our findings confirm the long-lasting mental health impact of SV and highlight the importance of specifically and routinely assessing SV in mental health care in old age as older victims do not seem to spontaneously disclose their experiences to mental health care professionals. In order to provide tailored care for older victims of SV, professionals working with old age psychiatry patients urgently need capacity building through training, screening tools and care procedures.

## Data Availability

By request from the corresponding author.

## Acknowledgements

The authors want to thank Lotte De Schrijver, Joke Depraetere, Adina Cismaru Inescu, and Laurent Nisen for their input during the questionnaire development. Also, we want to thank our co-interviewers Rosie De Vidts and Lisa Thibau for their time and dedication to this project. Many thanks to the staff of the Old Age Psychiatry Departments of AZ Sint-Jan Bruges-Ostend AV, Psychiatric Hospital Karus and Psychiatric Hospital Alexianen Zorggroep Tienen for their support during the data collection process. Finally, we would like to thank Dr. Howard Ryland for his help with the language editing.

## Appendices

### Appendix 1. Sample size calculation

|  |  |
| --- | --- |
| Formula |  |
| | $n = Z_{\alpha}^2 \left( \frac{P(1-P)}{e^2} \right)$ |
| Z-value for a 95% confidence interval ($Z_{\alpha}$) | 1.96 |
| Estimated proportion of sexual victimisation (P) | 0.10 |
| Margin of error (e) | 0.05 |
| Estimated sample size (n) | 138 |

### Appendix 2. Detailed outcome measurements sexual victimisation

#### Hands-off sexual victimisation (no physical contact)

- *Sexual staring:* Someone stared at me in a sexual way or looked at my intimate body parts (e.g., breasts, vagina, penis, anus) when I didn’t want it to happen.
- *Sexual innuendo:* Someone made teasing comments of a sexual nature about my body or appearance even though I didn’t want it to happen.
- *Showing sexual images:* Someone showed me sexual or obscene materials such as pictures, videos, directly or over the internet (including email, social networks and chat platforms) even though I didn’t want to look at them. This does not include mass mailings or spam.
- *Sexual calls or texts:* Someone made unwelcome sexual or obscene phone calls or texts to me.
- *Voyeurism:* I caught someone watching me, taking photos or filming me when I didn’t want it to happen while I was undressing, nude or having sex.
- *Distribution of sexual images:* Someone distributed naked pictures or videos of me directly or over the internet (including email, social networks and chat platforms) when I didn’t want it to happen.
- *Exhibitionism:* Someone showed their intimate body parts (e.g., breasts, vagina, penis, anus) to me in a sexual way and/or masturbated in front of me when I didn’t want to see it.
- *Forcing to show intimate body parts:* Someone made me show my intimate body parts (e.g., breasts, vagina, penis, anus) online or face-to-face when I didn’t want to do it.

#### Hands-on sexual victimisation

##### Sexual abuse (physical contact but no penetration)

- *Kissing:* Someone kissed me against my will.
- *Touching in care:* Someone touched my intimate body parts (e.g., breasts, vagina, penis, anus) during care against my will.
- *Fondling/rubbing:* Someone fondled or rubbed up against my intimate body parts (e.g., breasts, vagina, penis, anus) against my will.
- *Forced undressing:* Someone removed (some of) my clothes against my will.

##### Rape and attempted rape (physical contact with attempted or completed penetration)

- *Oral penetration:* Someone had oral sex with me or made me give oral sex against my will.
- *Attempt of oral penetration:* Someone tried, but did not succeed, to have oral sex with me or tried to make me give oral sex against my will.
- *Vaginal or anal penetration:* Someone put their penis, finger(s) or object(s) into my vagina or anus against my will.
- *Attempt of vaginal or anal penetration:* Someone tried, but did not succeed to put their penis, finger(s) or object(s) into my vagina or anus against my will.
- *Forcing to penetrate:* Someone made me put my penis, finger(s) or object(s) into their (or someone’s) vagina or anus against my will.

